# Return to play after treating acute muscle injuries in elite football players with a multimodal therapy approach that includes a specific protocol of (almost) daily radial extracorporeal shock wave therapy

**DOI:** 10.1101/2020.02.18.20024653

**Authors:** James P.M. Morgan, Mario Hamm, Christoph Schmitz, Matthias H. Brem

**Affiliations:** Extracorporeal Shock Wave Research Unit, Chair of Neuroanatomy, Institute of Anatomy, Faculty of Medicine, LMU Munich, Munich, Germany; Task force “Future of Professional Football”, DFL Deutsche Fussball Liga, Frankfurt, Germany; Curathleticum clinic, Nuremberg, Germany; Division of Trauma Surgery, Department of Surgery, Faculty of Medicine, University Hospital Erlangen, Friedrich-Alexander University Erlangen-Nuremberg, Erlangen, Germany

**Author notes:** **Correspondence** Dr. Christoph Schmitz, M.D., Extracorporeal Shock Wave Research, Unit, Chair of Neuroanatomy, Institute of Anatomy, Faculty of Medicine, LMU Munich, D-80336 Munich, Germany.

## Abstract

**Aim:** To compare lay-off times achieved by treating acute muscle injuries in elite football players with a multimodal therapy approach that includes a specific protocol of almost daily radial extracorporeal shock wave therapy (rESWT)) with corresponding data reported in the literature.

**Methods:** We performed a retrospective analysis of treatments and recovery times of muscle injuries suffered by the players of an elite football team competing in the first/second German Bundesliga during a previous season.

**Results:** A total of 20 acute muscle injuries were investigated in the aforementioned season, of which eight (40%) were diagnosed as type 1a/muscular tightness injuries, five (25%) as type 2b/muscle strain injuries, four (20%) as type 3a/partial muscle tear injuries and three (15%) as contusions. All injuries were treated with the previously mentioned multimodal therapy approach. Compared with data reported by Ekstrand et al. (Br J Sports Med 2013;47:769-774), lay-off times (median / mean) were shortened by 54% and 58% respectively in the case of type 1a injuries, by 50% and 55% respectively in the case of type 2b injuries as well as by 8% and 21% respectively in the case of type 3a injuries. No adverse reactions were observed.

**Conclusions:** Overall, the multimodal therapy approach investigated in this study is a safe and effective treatment approach for treating type 1a and 2b acute muscle injuries amongst elite football players and may help to prevent more severe, structural muscle injuries.

**What are the findings?:**

➢ By treating acute muscle injuries suffered by the players of an elite football team competing in the first/second German Bundesliga during a previous season with a multimodal therapy approach (comprising cryotherapy, compression, manual therapy, resistance/weight-training, a progressive physiotherapy exercise programme and a specific protocol of (almost) daily radial extracorporeal shock wave therapy (rESWT)) we achieved median and mean lay-off times after type 1a (muscular tightness/hypertonicity) and 2b (muscular strain injury) muscle injuries that were 50% shorter than corresponding data reported in the literature (Ekstrand et al., Brit J Sports Med 2013;47:769-774).
➢ The percentage of structural muscle injuries, specifically type 3 (partial muscle tear according to the Müller-Wohlfahrt/Munich muscle injury classification) and type 4 (complete muscle tear and/or avulsion injury involving the tendon) amongst this sample group of players (including injury types 1-4 as classified by Müller-Wohlfahrt et al; and excluding contusions) that occurred during the entire season comprised 23.5% of all the muscle injuries suffered. In comparison, the percentage of structural muscle injuries amongst similar sample groups of elite football players has been found to be considerably higher - in the data set reported by Ekstrand et al. (2013) higher grade structural muscle injuries amongst elite European football teams typically make up 66.9% of all muscle injuries suffered during the course of one season.

**How might it impact on clinical practice in the future?:** Our findings emphasise the effective use of a multimodal therapy approach (comprising cryotherapy, compression, manual therapy, resistance/weight-training, a progressive physiotherapy exercise programme and a specific protocol of (almost) daily rESWT for substantially shortening lay-off times associated with functional/ultrastructural muscle injuries and possibly for preventing more severe structural muscle injuries amongst sportspeople.

## INTRODUCTION

Muscle injuries are the most common injury in football.^1 2^ According to Müller-Wohlfahrt *et al*^3^, muscle injuries can be classified into indirect muscle injuries (comprising functional and structural injuries) and direct injuries (contusion, laceration) (table 1). Some authors proposed to classify functional injuries as ultrastructural injuries^4^.

**Table 1.**
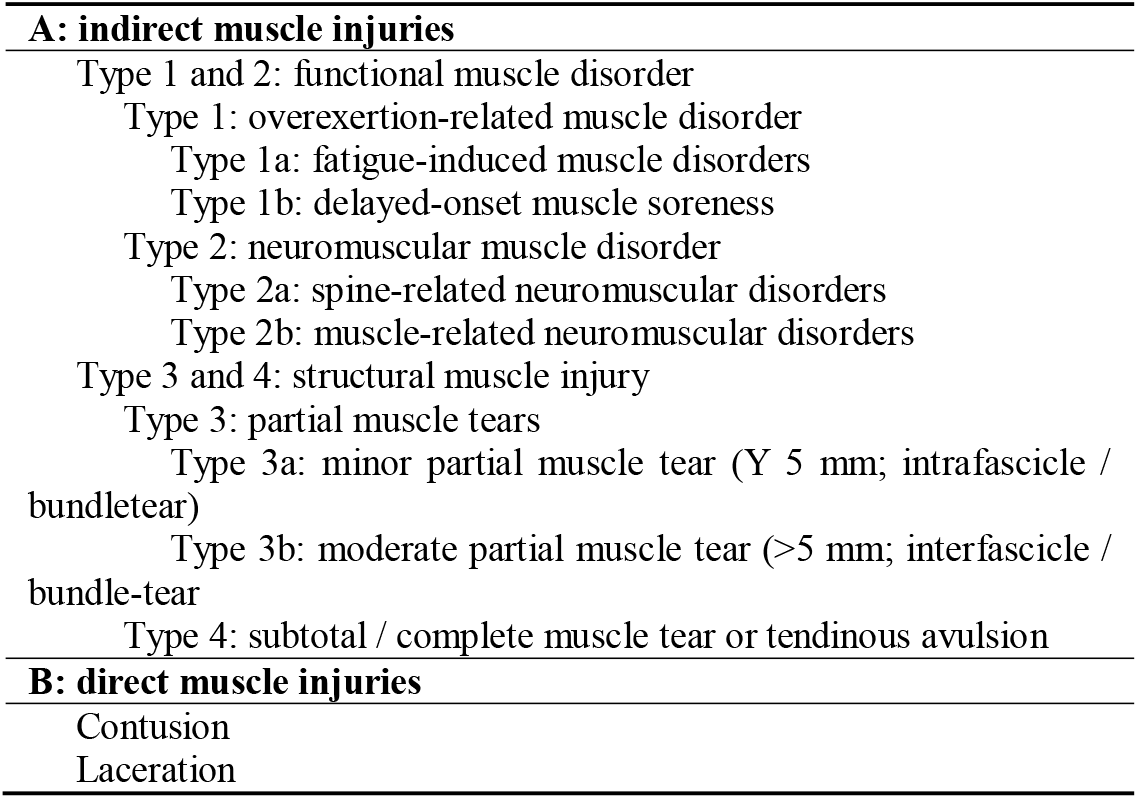
Classification of muscle injuries in sports (according to^3, 4^).

Recent approaches to improve therapy for acute muscle injuries have mainly focussed on type 3b injuries (structural injuries involving significant muscle tears).^5 6^ In contrast, the treatment of type 1a and 2b functional/ultrastructural muscle injuries as well as of type 3a structural muscle injuries (smaller partial muscle tears) has largely been neglected in the academic literature during the last few decades, although these injuries can also cause considerable lay-off times of two weeks or longer.^2 7^ In the guidelines for muscle injuries outlined by the Italian Society of Muscles, Ligaments and Tendons (ISMuLT), Maffulli *et al*^7^ recommended a multimodal therapy approach comprising RICE (rest, ice, compression, elevation), optimised loading, manual therapy, functional compression bandages, low-level laser therapy, pulsed ultrasound therapy, electroanalgesia, training and functional rehabilitation, without reference to specific evidence in the literature or modifying the treatment plan to cater for different types of injury severity.

During the past few years studies using animal subjects with acute muscle injuries and in vitro studies have shown that radial and focussed extracorporeal shock wave therapy (rESWT, fESWT) may be of benefit in treating acute muscle injuries.^8 9^ This form of treatment has already become well-established in successfully managing other pathologies of the musculoskeletal system, such as in the treatment of tendinopathies and fracture malunions.^10 11^ Radial and focussed extracorporeal shock waves (rESWs, fESWs) are single acoustic impulses which have an initial high positive peak pressure between 10 and 100 megapascals (MPa) that is reached in less than one microsecond (µs), followed by a low tensile amplitude of a few µs duration that can generate cavitation, and a short life cycle of approximately 10-20 μs.^12 13^ Due to these characteristics rESWs and fESWs fundamentally differ from therapeutic ultrasound. Focussed ESWs differ from rESWs in terms of how the shockwaves are generated. Focussed shock waves also differ in terms of their physical characteristics and with regards to the penetration depth of the shock waves into the tissue.^10 13-15^ Studies on rESWT/fESWT for the treatment of acute muscle injuries in elite football players (or other sportspeople) have not yet been published.

Based on anecdotal evidence we introduced rESWT (and to a much lesser extent, also fESWT) into our multimodal therapy approach for treating acute muscle injuries amongst elite football players (first/second German Bundesliga). Other components of this multimodal therapy approach included cryotherapy, compression, manual therapy, resistance/weight-training and a progressive physiotherapy exercise programme (in line with Maffulli *et al*^7^).

In this study we report our experience from the first entire season during which rESWT was applied. Our retrospective analysis was performed under the hypothesis that by integrating rESWT into our multimodal therapy approach, lay-off times may be shortened and a reduction of re-injury rates following acute functional/ultrastructural and structural muscle injuries amongst elite football players may be achieved.

## METHODS

We performed a retrospective analysis of treatments and time courses of all muscle injuries suffered by the players of an elite football team during a previous season (first/second German Bundesliga). This study was approved by the football club whose players were included in the study, and the local ethics board of Friedrich-Alexander University Erlangen-Nuremberg (Erlangen, Germany) (registration number 387_17 Bc). The players whose individual data and images are shown in figures 1 and 2 as well as in figure S1 and table S1 in Supplementary Information have explicitly granted their permission to publish these images. CS did not visit the football club during diagnosis and treatment sessions and only had access to fully anonymous data.

**Figure 1.**
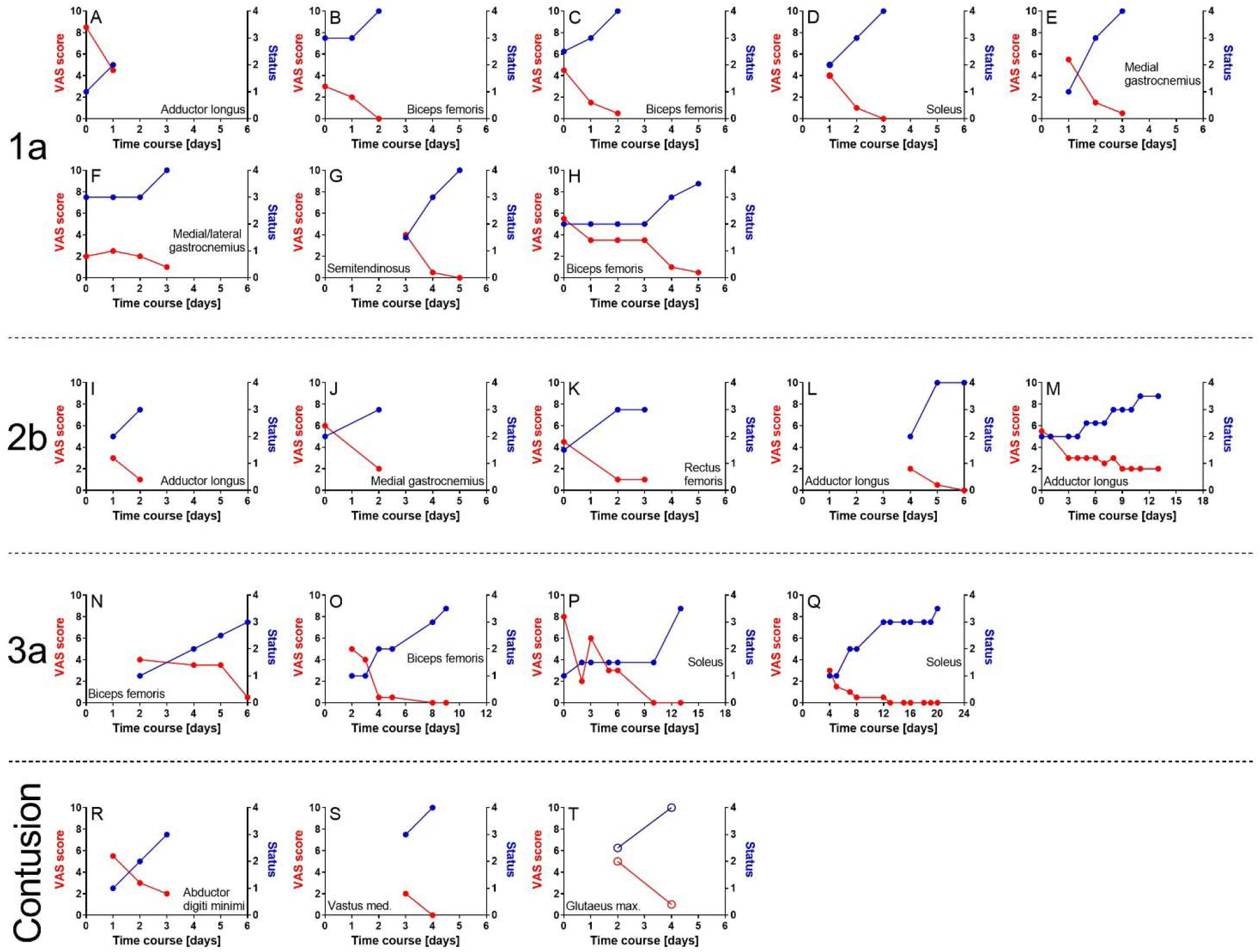
Time course of treatments of acute muscle injuries type 1a (A-H), 2b (I-M), 3a (N-Q) and contusions (R-T) suffered by the players of an elite football team during one of the previous seasons (first/second German Bundesliga), arranged in order of increasing lay-off times. VAS scores are marked by red dots (0 = no pain; 10 = maximal pain) and the player’s status by blue dots (1 = injured; 2 = rehabilitation; 3 = training; 4 = fully fit/return to play). Diagrams A-S represent treatments with radial extracorporeal shock wave therapy (rESWT), whereas Diagram T represents treatments with focused extracorporeal shock wave therapy (fESWT). In every case Day 0 was the day of injury; and a pair of red and blue dots indicate a single treatment session. When the player’s status was 4 on the day of the last treatment, return to play was achieved on this day. In contrast, when the player’s status was 3 on the day of the last treatment, return to play was achieved on the following day. Delays in commencing rESWT/fESWT were due to away games and travelling.

**Figure 2.**
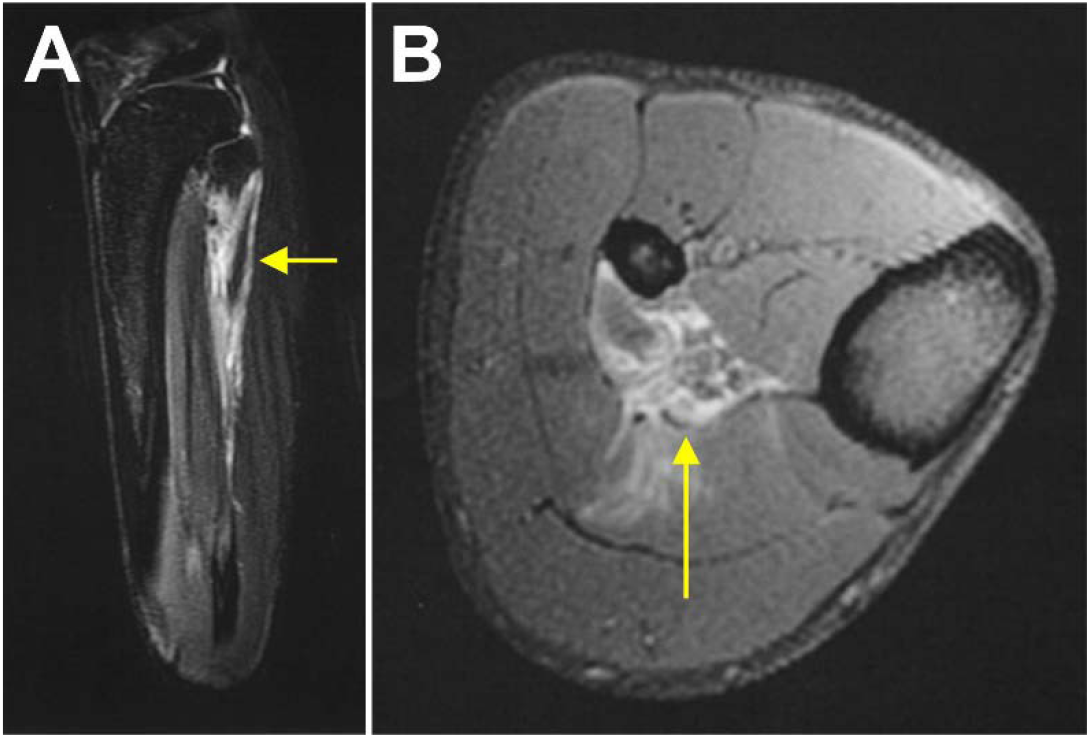
MRI images of the lower leg of an elite football player who was diagnosed with a minor partial muscle tear (type 3a) of the right soleus muscle (yellow arrows). The time course of the rehabilitation of this player is shown in figure 1Q; the detailed treatment protocol is summarised in table S1. Return to play was achieved on Day 22 after injury.

All players were male and aged between 18 and 35 years old. All other details (including the dates of injury, side of injury, the player’s position, whether an injured player was left-footed or right-footed, and whether an injury involved a player’s support leg or kicking leg) are subject to confidentiality in order to protect the identity of each player within this study.

The diagnosis of the acute muscle injuries within this study followed international guidelines and widely accepted clinical practice.^7^ In summary, the diagnosis of functional/ultrastructural injuries (type 1a and type 2b injuries respectively) was based on clinical examination. Specifically, functional tests such as hopping on one leg were used to provoke pain as well as to test the player’s readiness to commence (running) training; muscle length testing (stretch tests) and comparison with the non-injured limb, manual muscle strength testing and comparison with the non-injured limb, palpation of the injury site and comparison with the non-injured limb and a comprehensive subjective examination was undertaken. Pain was recorded using the Visual Analague Scale (VAS) during the clinical examination. The subjective examination included noting the mechanism of injury and the exact injury location as well as noting the nature or quality of the pain being experienced (tension, tightness, sharp pain, burning, etc). The diagnosis of type 2b, 3a and 3b injuries respectively was also performed by using the previously described clinical examination methods. In addition, ultrasound scans and magnetic resonance imaging (MRI) was used to differentiate between injury severity amongst these group of injuries. Contusions were diagnosed based on the patient’s history, clinical examination and ultrasound examination.

All acute muscle injuries were treated with a customised multimodal therapy approach comprising cryotherapy, compression, manual therapy, resistance/weight-training, a progressive physiotherapy exercise programme and ESWT (an example is provided in table S1 in Supplementary Information).

In 19 out of 20 cases rESWT was performed, using a Swiss DolorClast device (Electro Medical Systems, Nyon, Switzerland) equipped with an EvoBlue handpiece and 36-mm applicator. Radial ESWs were applied at 20 Hz. In the majority of cases rESWT was performed on a daily basis. The energy density of the rESWs was individually adjusted - so that the player reported some discomfort but did not experience pain during treatment, resulting in an air pressure of between 1.0 and 3.4 bar. A single treatment session consisted of between 6.000 and 12.000 rESWs being applied. Individual rESWT protocols are displayed in figure S1 in Supplementary Information. The decision to incorporate rESWT protocols into our treatment plan for muscle injuries was based on our promising earlier clinical experience and observations as well as positive subjective player information and feedback indicating that faster recovery times may be achievable in comparison to treatments of muscle injuries without the use of rESWT.

One case (contusion of the gluteus maximus muscle) was treated with fESWT, using a Swiss PiezoClast device (Electro Medical Systems) and 15-mm gel pad. Focussed ESWs were applied at 8 Hz; the positive energy density of the fESWs was 0.13 mJ/mm^2^ (energy level 10). A single treatment session consisted of between 2.500 and 3.000 fESWs being applied. The corresponding fESWT protocol is shown in figure S1Q in Supplementary Information.

In the vast majority of cases rESWT/fESWT was commenced on the day of the injury (9/20=45%), the day after the injury (4/20=25%) or two days after the injury (3/20=15%). Delays in starting with rESWT/fESWT were due to away games or travelling and as a result not having immediate access to shock wave therapy.

There were no contraindications to rESWT amongst the group of players that were treated. Contraindications to rESWT include: local steroid injections during the last six weeks before rESWT/fESWT, infection or tumor at the site of rESWT/fESWT application, serious blood dyscrasia, blood-clotting disorders (including local thrombosis) and treatment with oral anticoagulants.

“Return-to-sport” status was defined as being once the player was able to fully participate in regular team training including contact training; and was fully available for selection for matches in the first/second Bundesliga.

## RESULTS

A total of 20 acute muscle injuries occurred during the investigated season and were treated with the aforementioned approach, of which eight (40%) were diagnosed as type 1a injuries, five (25%) as type 2b injuries, four (20%) as type 3a injuries and three (15%) as contusions (figure 1). Accordingly, 13/17=76% of the injuries that were not contusions were functional/ultrastructural injuries (types 1a and 2b), and 4/17=24% were structural injuries (type 3a). There were no type 3b or type 4 injuries suffered during the investigated season.

Amongst the 17 injuries that were not classified as contusions, one (6%) occurred in the muscles in the anterior compartment of the thigh, four (24%) in the muscles in the medial compartment of the thigh, six (35%) in the posterior compartment of the thigh, and six (35%) in the posterior compartment of the leg.

Injuries occurred during the entire season. There was no difference in the number of injuries suffered at the beginning/first half of the season compared to the injuries suffered in the second half of the season after the winter break. All of the structural injuries (type 3a) occurred several months after the start of the season. The re-injury rate (defined as being an injury involving the same muscle and having the same severity as the initial injury occurring within two months after a player’s return-to-sport following the initial injury^2^) was 1/8=12.5% for type 1a injuries, and was zero in the case of type 2b, 3a injuries and contusions respectively. None of the type 2b injuries that were diagnosed occurred following a previously suffered type 1a injury during the investigated season, and none of the type 3a injuries occurred following a previously suffered type 1a or 2b injury during the investigated season.

Return-to-play was achieved after 3 / 3.3 / 2-6 days (median / mean / range) respectively following type 1a injuries, after 4 / 6.2 / 3-14 days respectively following type 2b injuries, after 12 / 13 / 7-22 days respectively following type 3a injuries and after 4 / 4 / 4 days respectively following contusions.

MRI pictures of the lower leg of a player who was diagnosed with a minor partial muscle tear (type 3a) of the soleus muscle are shown in figure 2. In this case rESWT was performed on days 4, 5, 7, 8, 12, 13, 15, 16 and 18-20 respectively following injury (figure 1Q). The detailed treatment protocol of this case is summarized in table S1 in Supplementary Information. Return-to-play was achieved on day 22 post-injury.

No adverse reactions to treatment were observed, except occasional, temporary reddening of the skin at the treatment site that disappeared within 24 hours following rESWT.

All of the players who were treated for muscle injuries complied fully with the treatment protocol incorporating ESWT. No player declined being treated with ESWT neither during the initial treatment nor during the follow-up treatments. Hence, by comparing the use of our treatment protocol for muscle injuries with other treatment protocols and the differences in lay-off times respectively, one can conclude that non-compliance did not affect the treatment effect of our retrospective analysis.

## DISCUSSION

This is the first report concerning the course of recovery times following acute muscle injuries suffered by the players of an elite football team (competing in the first/second German Bundesliga) during an entire season that incorporated rESWT/fESWT into the treatment protocols of these injuries. A number of relevant conclusions can be drawn from these results (with reference to rESWT because fESWT was only used in one single case).

Firstly, integrating rESWT into a multimodal therapy approach for treating acute muscle injuries as outlined in this study is safe. There were no adverse reactions or complications to treatment observed apart from occasional, temporary reddening of the skin at the treatment site that disappeared within 24 hours following rESWT. Moreover, there were no observed cases of local hematoma following the application of rESWs - as has been previously reported in the literature following the application of rESWs for the treatment of the gluteus maximus muscle with a rESWT device that differed from the one that was used in this study^16^. In addition, none of the players that were treated suffered from myositis ossificans (MO) - a proliferative mesenchymal response following soft tissue trauma that causes localised ossification.^17^ This is important to note because, at first glance, exposure of injured muscles to rESWs could be considered inappropriate physiotherapy.^18^ Walczak *et al*^17^ hypothesized (with reference to a study by Medici *et al*^19^) that the development of MO may depend on a process called endothelial-mesenchymal transition. According to this hypothesis, skeletal muscle injury may induce a local inflammatory cascade, which leads to the release of bone morphogenetic protein (BMP)-2, BMP-4 and transforming growth factor (TGF). These cytokines may act on vascular endothelial cells and induce endothelial-mesenchymal transition. As a result, endothelial-derived mesenchymal stem cells may differentiate into osteoblasts and chondrocytes when exposed to an inflammatory-rich environment.^17.^ Accordingly, it is important to note that exposure of tissue to ESWs (or, more specifically, exposure to *f*ESWs; because related studies on rESWs have not yet been published) can increase local concentrations of BMP-2, BMP-4 and TGF-ß1.^20,21^ If this is the case it would clearly be an argument against using rESWT/fESWT in the treatment of acute muscle injuries. However, it is important to keep in mind that the aforementioned hypothesis of endothelial-mesenchymal transition in the pathophysiology of MO is based on observations of a rare disease called fibrodysplasia ossificans progressiva^19^ rather than on traumatic MO. A number of studies on human heterotopic ossification and related mouse models have demonstrated that bone marrow-derived osteoblast progenitor cells in circulating blood may contribute to the formation of heterotopic bone.^22-24^ Moreover, a recent study involving a parabiosis model (i.e. two mice with shared circulation, in which the cells of one mouse can be labelled and detected in the other mouse when released into the blood stream) has demonstrated that these bone marrow-derived osteoblast progenitor cells do not contribute to early stage development of heterotopic ossification.^25^ The authors of this study argued that a timely intervention that influences the molecular and cellular processes that contribute to the recruitment and/or differentiation of the circulating cell populations that participate in heterotopic ossification may block the initiation of the latter and ultimately prevent the transition to definitive bone.^25^ We believe that this may be achieved by the very early treatment of acute muscle injuries with rESWT in this study. Further studies on animal models are required to test this hypothesis. It is worth noting that in this study commencing rESWT immediately following type 1a and 2b injuries did not lead to inferior results when compared with starting treatment with rESWT on days 1, 3 or 4 post-injury (delay in commencing treatment was due to some injuries being suffered during away games that required travelling after the games and as a result there being no immediate access to rESWT).

Secondly, integrating rESWT into a multimodal therapy approach for the treatment of acute muscle injuries as performed in this study may shorten lay-off times for elite football players (as well as for other sportspeople) compared to other therapy approaches. This conclusion is based on a comparison of the results of this study with data that was obtained prospectively from 31 European elite male football teams during the 2011/2012 season (table 2).^2^

**Table 2.** Comparison of lay-off times after acute muscle injuries in elite football players reported in this study as well as by Ekstrand *et al*^2^.

|  | Type 1a |  |  | Type 2b |  |  | Type 3a |  |  |  |
| --- | --- | --- | --- | --- | --- | --- | --- | --- | --- | --- |
|  | X<sup>a</sup> | E<sup>b</sup> | D<sup>c</sup> | X<sup>a</sup> | E<sup>b</sup> | D<sup>c</sup> | X<sup>a</sup> | E (3a)<sup>d</sup> | D<sup>c</sup> | E (3b)<sup>e</sup> |
| n | 8 | 82 |  | 5 | 22 |  | 4 (3) | 192 |  | 66 |
| Median | 3 | 6.5 | **-54%** | 4 | 8 | **-50%** | 12 (10) | 13 | **-8% (-23%)** | 32 |
| Mean | 3.3 | 7.9 | **-58%** | 6.2 | 13.8 | **-55%** | 13 (10.3) | 16.4 | **-21% (-37%)** | 37.9 |
| SD | 1.5 | 7.4 |  | 4.7 | 20.6 |  | 6.1 (3.5) | 17.1 |  | 24.0 |
| Min | 2 | 1 |  | 3 | 2 |  | 7 | 2 |  | 8 |
| Max | 6 | 58 |  | 14 | 100 |  | 14 | 132 |  | 156 |

<sup>a</sup> Results of this study.<sup>b</sup> Results reported by Ekstrand *et al*<sup>2</sup>.<sup>c</sup> Percentage difference<sup>d</sup> Results reported by Ekstrand *et al*<sup>2</sup> for minor partial muscle tears (i.e., Type 3a).<sup>e</sup> Results reported by Ekstrand *et al*<sup>2</sup> for moderate partial muscle tears (i.e., Type 3b).

A direct comparison with the data reported by Ekstrand *et al*^2^ shows that the median / mean lay-off times in this study were shortened by 54% / 58% respectively in the case of type 1a injuries, by 50% / 55% respectively in the case of type 2b injuries, and by 8% / 21% respectively in the case of type 3a injuries. Arguably, the type 3a structural injury shown in figure 2 could also be considered to be a more severe type 3b injury rather than a type 3a injury which would mean that the median / mean injury lay-off times in this study for 3a injuries would then be further shortened by 23% / 37% respectively compared to the data reported by Ekstrand *et al*^2^. Furthermore, in recently published guidelines for muscle injuries, Maffulli *et al*^7^ consider normal lay-off times to be 5-15 days for functional/ultrastructural injuries, 15-18 days for type 3a injuries and 25-35 days for type 3b injuries. The lay-off times reported in this study were substantially shorter than the aforementioned typical lay-off times noted by Maffulli *et al*.^7^ At this stage it is difficult to ascertain to which extent the application of rESWs actually contributed to the shorter lay-off times in this study compared to the data reported by Ekstrand *et al*.^2^ This is partly due to the fact that no treatment protocols were outlined in the latter study. Nevertheless, several factors strengthen the argumentation that the application of rESWs may be helpful as an adjunct form of therapy for reducing lay-off times following muscle injuries. Specifically: (i) the study by Maffulli *et al*.^7^ was based on data that was obtained from European male elite football teams and, therefore, from a population that is very similar to the population investigated in this study; (ii) the key molecular and cellular mechanisms of action that rESWs/fESWs have on muscles (outlined below) were not yet known in 2011/2012; (iii) Maffulli *et al*^7^ did not mention rESWT/fESWT in their guidelines for muscle injuries; and (iv) our multimodal therapy approach (excluding rESWT/fESWT) is in line with the guidelines published by Maffulli *et al*.^7.^ With this in mind it is reasonable to assume that the therapy approaches used in 2011/2012 by the teams who were investigated by Ekstrand *et al*^2^ were broadly comparable to the multimodal therapy approach (excluding rESWT/fESWT) used in this study. Hence the application of rESWs may have substantially contributed to the shorter lay-off times found in this study compared to the data reported by Ekstrand *et al*.^2^ Further studies are required to test this hypothesis.

In addition, the total re-injury rates of 1/13 (8%) observed amongst functional/ultrastructural muscle injuries and 0/4 (0%) observed amongst the structural muscle injuries in this study were lower than corresponding data reported by Ekstrand *et al*^2^ (12% was observed amongst functional/ultrastructural injuries and 13% observed amongst structural injuries respectively). Based on these results we argue that the integration of rESWT into a multimodal therapy approach for the treatment of acute muscle injuries as outlined in this study may not only help to reduce lay-off times but may also help in the prevention of muscle re-injury amongst athletes. However, due to the low number of structural muscle injuries included in this study, further studies are required to test this hypothesis amongst a larger sample group.

In addition, no type 2b injuries occurred in this study following a type 1a injury and no structural injury occurred following a functional/ultrastructural injury. In contrast, Ekstrand *et al*^2^ reported in their study, that 5% of the initial functional/ultrastructural injuries developed into secondary structural injuries within two months of the primary injury. However, they did not report how many type 2b injuries occurred following an earlier type 1a injury during the investigated season, nor did they comment on how many structural injuries occurred following previously suffered functional/ultrastructural injuries during the entire investigated season (i.e. not only including the initial two months, or “re-injury” period following the primary injury). It is important to note that the number of functional/ultrastructural compared to structural acute muscle injuries was 13 as opposed to four. Hence there were considerably more functional/ultrastructural injuries included in this study, but this number was 130 as opposed to 263 (i.e. more structural injuries) in the study by Ekstrand *et al*.^2^ Of course many other factors may have contributed to this finding - such as differences in the management of training load upon returning to team training, and other prevention strategies used such as optimising nutrition and using screening protocols for detecting early warning signs that may allow sport medical staff to help “forecast” an impending muscle injury^26-28^. However, one cannot rule out that the integration of rESWT into a multimodal therapy approach for the treatment of acute muscle injuries as performed in this study may also contribute to the prevention of structural muscle injuries in athletes.

This study is an audit of retrospectively collected data, and therefore has a number of inherent limitations. Firstly, there was no control group, which meant that the outcomes of the players of the same elite football team that suffered from acute muscle injuries and were treated with the same multimodal therapy approach (following diagnosis and treatment by the same doctors and physiotherapists) could not be directly compared with those that were treated without rESWT/fESWT. The reason for this was that the evidence that was available at the time when we decided to integrate rESWT/fESWT into our therapy approach was not sufficient enough to gain approval from the club for a possible prospective study with a control group. Paradoxically, the promising results of this study - in particular the comparison to the data reported by Ekstrand *et al*^2^, may now also cause difficulties in gaining approval from the club for a corresponding prospective study with a control group - as the club’s interest obviously lies in reducing lay-off times following injuries to a minimum. Moreover, gaining approval for a corresponding randomized controlled trial (RCT) from the players themselves may also prove to be more difficult following the initial good results obtained from using rESWT/fESWT in this study. Perhaps it is more realistic to expect that future RCTs be performed on recreational athletes, as in the case of one current study that relates to the treatment of acute type 3b hamstring muscle injuries.^15^

Secondly, with the exception of one case (contusion) only rESWT was investigated in this study. One may also argue that fESWT may also be effective in the management of acute muscle injuries amongst athletes, but obviously there is not enough data to support this hypothesis. Our decision to focus on rESWT rather than on fESWT was based on the fact that the data relating to treatment of other pathologies of the musculoskeletal system (tendinopathies and fracture malunions of superficial bones) do not support the hypothesis that fESWT is superior to rESWT,^10,11^ In addition, the use of fESWT is restricted to physicians in many countries (as is the case in Germany where physiotherapists and chiropractors who have trained in Germany are not legally entitled to use fESWT). Moreover, the International Society for Medical Shockwave Treatment (ISMST) has recommended that only a qualified physician (certified by National or International Societies) may use fESWT in their latest Consensus Statement regarding ESWT indications and contraindications.^29^ However, in many elite football clubs (and training centres involving other sporting codes) a medical doctor may not be available to perform treatment on a daily basis. This is also the reason why a RCT on acute type 3b hamstring muscle injuries currently being undertaken is based on rESWT rather than on fESWT.^15^

Thirdly, only one rESWT protocol was applied in this study, and this protocol differed considerably from other published rESWT protocols used in the treatments of tendinopathies^10^. Specifically, differences regarding the timing with regards to commencing rESWT (immediately after the injury in this study compared with starting treatment six or more months after initial diagnosis and following unsuccessful treatment involving other conservative modalities)^30-32^, differences in the time interval between treatment sessions - in most cases daily treatment sessions were used in this study as compared to one treatment session per week used in other studies^10^, as well as differences in the number of rESWs applied per treatment session (6.000-12.000 in this study as opposed to treatments averaging 2.000)^10^. As already mentioned above, our motivation to use this particular rESWT protocol was based on our (and the players’) clinical experience and observations that players achieved faster recovery times than in our earlier treatments of muscle injuries that did not include using rESWT (which has been confirmed by comparing the data set in this study with the data by Ekstrand *et al*^2^). Nevertheless, we cannot rule out the possibility that a time interval of two days between treatment sessions, the application of fewer than 6.000 rESWs per treatment session and/or application of rESWs with lower energy densities than those applied in this study may lead to the same results (reduced lay-off times, reduced re-injury rates, etc.) in the treatment of acute muscle injuries amongst elite football players as reported in this study. This may be addressed in follow-up studies.

Finally, the precise molecular and cellular mechanisms that may have contributed to the outcomes of this study as a result of incorporating rESWT into our treatment protocol remain unknown - there were no muscle biopsies or blood samples taken in this study. In addition, there have been no studies published using animal models with similar muscle injuries as those that were investigated in this study in which rESWT was applied in conjunction with other treatment modalities that would mimic our multimodal therapy approach. The molecular and cellular mechanisms of rESWT include: (i) the depletion of presynaptic substance P from C-fibers^33^ leading to a reduction in the sensation of pain and blockage of neurogenic inflammation;^34^ (ii) muscular relaxation possibly caused by the mechanical separation of actin and myosin filaments and/or transient dysfunction of nerve conduction at neuromuscular junctions^35,36^; (iii) stimulation of tissue remodelling by promoting inflammatory and catabolic processes that are associated with the removal of damaged matrix constituents^37^; (iv) enhanced proliferation and differentiation rates and modulation of gene expression of muscle satellite cells^8,9^; (v) stimulation of fibroblast proliferation^13^; (vi) improved fascial gliding within the surrounding tissues due to increased lubricin expression^38^; and (vii) functional angiogenesis / improved blood circulation^39,40^. Many of these mechanisms may also have a positive carryover on the other treatment modalities used in our multimodal therapy approach. For example, pain reduction may allow manual therapy to be carried out more effectively, and improved fascial movement may contribute to better performance during training. These assumptions need to be supported by further studies.

## CONCLUSIONS

This study suggests that integrating rESWT into a multimodal therapy approach for the treatment of acute type 1a, 2b, 3a muscle injuries and contusions is safe and effective, and leads to shortened lay-off times and reduced re-injury rates amongst elite football players, without causing any adverse effects. Clinicians should consider rESWT in the management of acute type 1a, 2b 3a muscle injuries and contusions in athletes and sportspeople.

## Data Availability

Due to confidentiality reasons, there are no data that can be shared except for those that are provided in the manuscript.

## Contributors

JPMM performed all treatments and documented treatment outcome, reviewed and revised the manuscript. CS drafted the manuscript. MH and MHB reviewed and revised the manuscript. JPMM, MH, CS and MHB designed the study, analysed and interpreted the data.

## Patient consent

All players whose individual healing process and course of treatment are shown in figures 1 and S1 as well as the player whose individual MRI scans are shown in figure 2 have explicitely granted permission to publish these data and images.

## Ethics approval

This study was approved by the local ethics board of Friedrich-Alexander University Erlangen-Nuremberg (Erlangen, Germany).

## Provenance and peer review

Not commissioned; externally peer reviewed.

## Data sharing statement

Due to confidentiality reasons, there are no data that can be shared.

---

**Figure S1.**
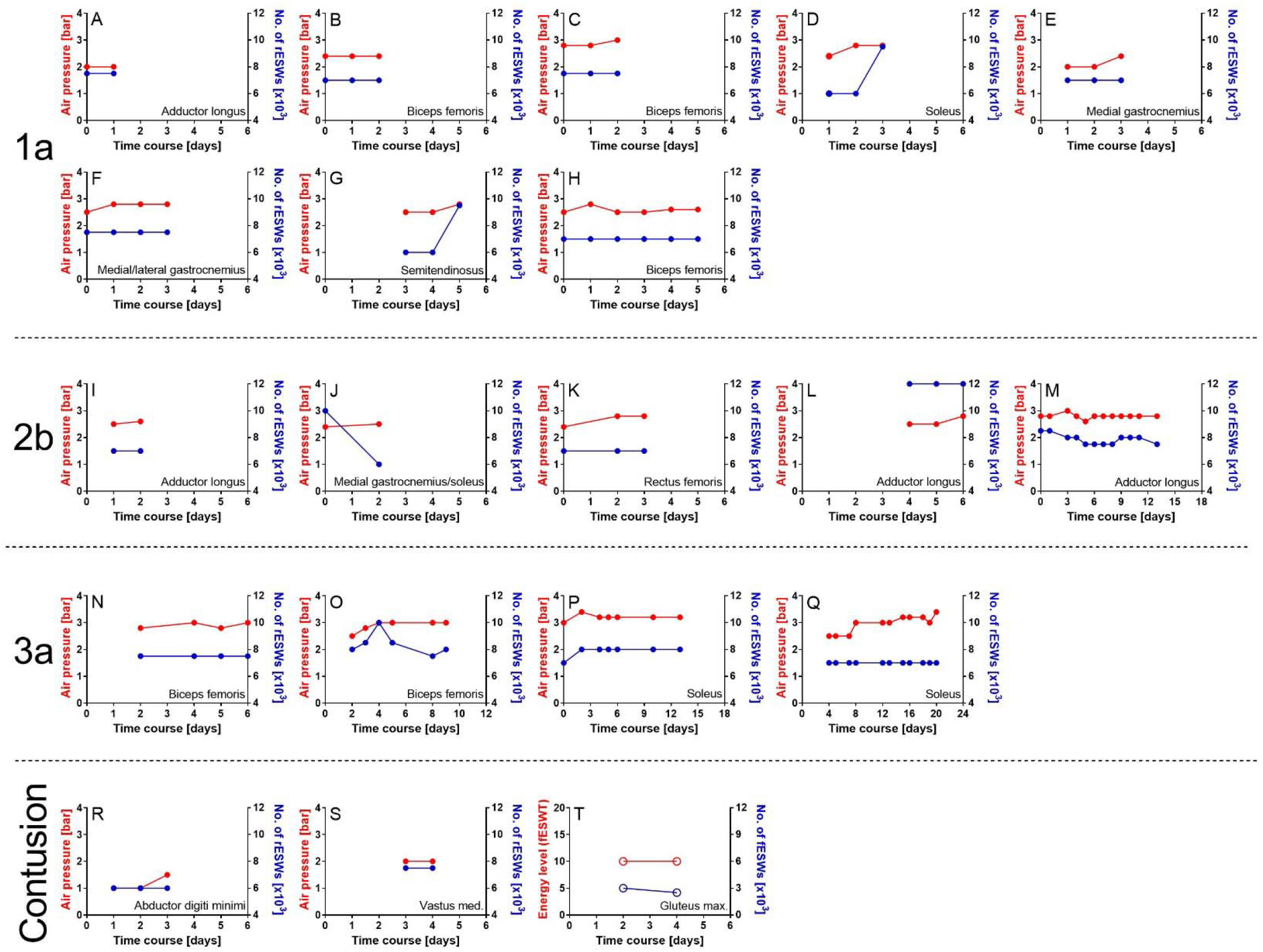
Protocols of radial extracorporeal shock wave therapy (rESWT) (A-S) or focused extracorporeal shock wave therapy (fESWT) (T) of all acute muscle injuries type 1a (A-H), 2b (I-M), 3a (N-Q) and contusions (R-T) suffered by the players of an elite football team during one of the previous seasons (first/second German Bundesliga), arranged in order of increasing lay-off times. All rESWT treatments were performed with a Swiss DolorClast device (Electro Medical Systems, Nyon, Switzerland) equipped with EvoBlue handpiece and 36-mm applicator; radial extracorporeal shock waves (rESWs) were applied at 20 Hz. Air pressure data is marked by red dots (between 2 and 3.5 bar) and the number of rESWs per treatment session by blue dots (between 6.000 and 12,000 per treatment session). The fESWT treatment shown in T was performed with a Swiss PiezoClast (Electro Medical Systems) and 15-mm gel pad; focused extracorporeal shock waves (fESWs) were applied at 8 Hz. Energy level data are marked by red dots (Level 10) and the number of fESWs per treatment session by blue dots (between 2.500 and 3.000 per treatment). In each case Day 0 was the day of injury. Every pair of red and blue dots indicate a single treatment session. When the player’s status was 4 on the day of the last treatment, return to play was achieved on this day. In contrast, when the player’s status was 3 on the day of the last treatment, return to play was achieved on the following day. Delays in starting with rESWT were due to away games and travelling.

**Table S1.** Treatment protocol of a player who was diagnosed with a partial muscle tear (type 3a) of the right soleus muscle (c.f. figures 1Q and 2 in the main text as well as figure S1Q in Supplementary information).

| Day | Physiotherapy | Rehabilitation | rESWT<sup>a</sup> |
| --- | --- | --- | --- |
| 1 | Cryo<sup>b</sup>, Comp<sup>c</sup> |  |  |
| 2 | Cryo<sup>b</sup>, Comp<sup>c</sup> |  |  |
| 3 | Cryo<sup>b</sup>, Comp<sup>c</sup> |  |  |
| 4 | Cryo<sup>b</sup>, Comp<sup>c</sup>, MT<sup>d</sup> | Swimming | 7000 / 2.5 |
| 5 | Cryo<sup>b</sup>, Comp<sup>c</sup>, MT<sup>d</sup> | Swimming | 7000 / 2.5 |
| 6 | Cryo<sup>b</sup>, Comp<sup>c</sup>, MT<sup>d</sup> | Swimming, R/W/P<sup>e</sup> triceps surae (TheraBand) |  |
| 7 | Cryo<sup>b</sup>, Comp<sup>c</sup>, MT<sup>d</sup> |  | 7000 / 2.5 |
| 8 | MT<sup>d</sup> | Bicycling, R/W/P<sup>e</sup> triceps surae (TheraBand) | 7000 / 3.0 |
| 9 | MT<sup>d</sup> | Bicycling, R/W/P<sup>e</sup> triceps surae (TheraBand) |  |
| 10 | MT<sup>d</sup> | Bicycling, R/W/P<sup>e</sup> triceps surae (training device) |  |
| 11 | MT<sup>d</sup> | Bicycling |  |
| 12 | MT<sup>d</sup> | Bicycling, R/W/P<sup>e</sup> triceps surae (training device) | 7000 / 3.0 |
| 13 | MT<sup>d</sup> | Running (10 × 1 min running followed by 1 min walking) | 7000 / 3.0 |
| 14 |  | Running (10 × 2 min running followed by 1 min walking) |  |
| 15 | MT<sup>d</sup> | R/W/P<sup>e</sup> triceps surae (training device) | 7000 / 3.2 |
| 16 | MT<sup>d</sup> | Running (2 × 10 min) | 7000 / 3.2 |
| 17 | MT<sup>d</sup> | R/W/P<sup>e</sup> triceps surae (training device) |  |
| 18 | MT<sup>d</sup> |  | 7000 / 3.2 |
| 19 | MT | R/W/P<sup>e</sup> triceps surae (training device), running (20 min) | 7000 / 3.0 |
| 20 | MT<sup>d</sup> | Running (30 min) | 7000 / 3.4 |
| 21 |  | Running (40 min) incl. 4 × 3 min at 80% lactate threshold |  |

<sup>a</sup> Radial extracorporeal shock wave therapy (Swiss DolorClast [Electro Medical Systems, Nyon, Switzerland] with EvoBlue handpiece and 36-mm applicator): number of radial extracorporeal shock waves per session / air pressure (bar).
<sup>b</sup> Cryotherapy.
<sup>c</sup> Compression.
<sup>d</sup> Manual therapy.
<sup>e</sup> Resistance/weight-training/progressive physiotherapy exercise programme.

